# Harnessing ChatGPT and GPT-4 for Evaluating the Rheumatology Questions of the Spanish Access Exam to Specialized Medical Training

**DOI:** 10.1101/2023.07.21.23292821

**Authors:** Alfredo Madrid-García, Zulema Rosales-Rosado, Dalifer Freites-Nuñez, Inés Pérez-Sancristobal, Esperanza Pato-Cour, Chamaida Plasencia-Rodríguez, Luis Cabeza-Osorio, Leticia León-Mateos, Lydia Abasolo-Alcázar, Benjamín Fernández-Gutiérrez, Luis Rodríguez-Rodríguez

**Affiliations:** Grupo de Patología Musculoesquelética. Hospital Clínico San Carlos. Instituto de Investigación Sanitaria San Carlos (IdISSC), Prof. Martin Lagos s/n, Madrid, 28040, Spain; Medicina Interna. Hospital Universitario del Henares, Avenida de Marie Curie, 0, 28822, Coslada, Madrid, Madrid, 28822, Spain; Facultad de Medicina. Universidad Francisco de Vitoria, Carretera Pozuelo, km 1800, 28223, Majadahonda, Madrid, Madrid, 28223, Spain; Reumatología. Hospital Universitario la Paz-IdiPaz, Paseo de la Castellana, 261, 28046 Madrid, Madrid, 28046, Spain

**Author notes:** Corresponding author (A. Madrid-García). The authors declare there are no competing interests. First author (Alfredo Madrid-García). These authors contributed equally (Zulema Rosales-Rosado, Dalifer Freites-Nuñez, Inés Pérez-Sancristobal, Esperanza Pato-Cour, Chamaida Plasencia-Rodríguez, Luis Cabeza-Osorio).

**Keywords:** Artificial intelligence, Chatbot, ChatGPT, GPT-4, Transformer, Large language model, Education, Rheumatology, Natural language processing

## Abstract

The emergence of Large Language Models (LLM) with remarkable performance such as ChatGPT and GPT-4, has led to an unprecedented uptake in the population. One of their most promising and studied applications concerns education due to their ability to understand and generate human-like text, creating a multitude of opportunities for enhancing educational practices and outcomes. The objective of this study is two-fold: to assess the accuracy of ChatGPT/GPT-4 in answering rheumatology questions from the access exam to specialized medical training in Spain (MIR), and to evaluate the medical reasoning followed by these LLM to answer those questions. A dataset, RheumaMIR, of 145 rheumatology-related questions, extracted from the exams held between 2010 and 2023, was created for that purpose, used as a prompt for the LLM, and was publicly distributed. Six rheumatologists with clinical and teaching experience evaluated the clinical reasoning of the chatbots using a 5-point Likert scale and their degree of agreement was analyzed. The association between variables that could influence the models’ accuracy (i.e., year of the exam question, disease addressed, type of question and genre) was studied. ChatGPT demonstrated a high level of performance in both accuracy, 66.43%, and clinical reasoning, median (Q1-Q3), 4.5 (2.33-4.67). However, GPT-4 showed better performance with an accuracy score of 93.71% and a median clinical reasoning value of 4.67 (4.5-4.83). These findings suggest that LLM may serve as valuable tools in rheumatology education, aiding in exam preparation and supplementing traditional teaching methods.

**What is already known on this topic:** Large Language Models have demonstrated remarkable performance when presented with medical exam questions. However, no study has evaluated their clinical reasoning in the rheumatology field.

**What this study adds:** This is the first study to evaluate the accuracy and clinical reasoning of ChatGPT and GPT-4 when rheumatology questions from an official access exam to specialized medical training are used as prompts.

**How this study might affect research, practice or policy?:** This study highlights the usefulness of two Large Language Models, ChatGPT and GPT-4, in the training of medical students in the field of rheumatology.

**Highlights:**

- ChatGPT showed an accuracy of 66.43% in answering MIR questions, while GPT-4 exhibits a significantly higher proficiency with an accuracy of 93.71%.
- The median (Q1-Q3) value of the average score for the clinical reasoning of GPT-4 was 4.67 (4.5-4.83), while for ChatGPT was 4.5 (2.33-4.67).

## 1. Introduction

The emergence of Large Language Models (LLM) with remarkable performance such as ChatGPT, has led to an unprecedented uptake in the population, being the fastest-growing application in history [1], and with the potential to transform various domains, including medicine [2]. A clear example of this burgeoning interest is the notable surge in scientific research focusing on ChatGPT’s role within the medical field. From January 1^st^, 2023 until July 10^th^, 2023, there have been over 768 publications indexed in PubMed, that contain the string *ChatGPT* and that delve into various aspects of this Generative Pre-trained Transformer (GPT) chatbot. Some of the topics of such publications include the potential impact of ChatGPT on clinical and translational medicine [3], clinical pharmacology [4], scientific communication [5] and medical writing [6], medical evidence summarization [7], patient question-answering [8], public health [9], ethical and legal [10], health policy making [11], plastic and colorectal surgery [12, 13], doctor-patient communication [9], or drug-drug interaction and explainability [14].

In rheumatology, the use of AI and LLM, such chatbots, is gaining relevance [15]. A query conducted on July 10^th^ in PubMed, *“Rheumatology AND ChatGPT”* produced twelve results. For instance, in [16], the authors asked ChatGPT to draft an editorial about artificial intelligence (AI) potentially replacing rheumatologists in editorial writing and discussed the ethical aspects and the future role of rheumatologists. In [17], the authors discussed the role of ChatGPT as an author, and the inherent terms associated with the concept of authorship such as accountability, bias, accuracy, and responsibility. Eventually, they considered refusing to recognize ChatGPT as an author, in accordance also to *Nature* announcement [18]. For their part, the authors in [19] provided an overview of the potential use of ChatGPT as a rheumatologist, its capabilities and risks. They concluded that although chatbots will not be able to make clinical decisions for now; they will be able, for example, to design study protocols, streamline information accessibility and contribute to time-saving. A similar correspondence article highlighted five areas in which ChatGPT has the potential to assist in rheumatology care delivery, to note: patient drug education, medical imaging reporting, medical note-keeping for outpatient consultations, medical education and training and clinical audit and research. Finally, the authors in [20] used ChatGPT as an assistant tool to aid in writing a case report of a systemic lupus erythematosus patient.

One of the most promising and studied applications for tools like ChatGPT lies within the realm of education. The ability of these language models to understand and generate human-like text creates a multitude of opportunities for enhancing educational practices and outcomes [21, 22, 23, 24, 25]. Within this field, one of the most extensively studied applications of ChatGPT is its ability and reasoning in answering medical examination questions. One of the most prominent studies was [26]. This study demonstrated how ChatGPT performed at or near the passing threshold for all three tests of the United States Medical Licensing Exam despite not having undergone any specialized training or reinforcement. However, many similar studies have emerged in recent months.

Given the range of studies that have subjected ChatGPT to different medical exam questions (e.g., multiple-choice, open-ended), the disparity in the results obtained, and the proficiency of ChatGPT in understanding context and generating detailed responses, we hypothesize that these tools could aid in the learning process of medical students, particularly in the study of rheumatology and musculoskeletal diseases.

Therefore, the objective of this study is two-fold:

- First, we seek to ascertain the accuracy of ChatGPT/GPT-4 in answering rheumatology questions from the access exam to specialized medical training in Spain, Médico Interno Residente (MIR).
- Secondly, we aim to evaluate the clinical reasoning followed by ChatGPT/GPT-4 in answering those multiple-choice questions.

## 2. Materials and Methods

### 2.1. ChatGPT and GPT-4 as Large Language Models

ChatGPT, an iteration of the Generative Pre-trained Transformer (GPT) model, is an AI chatbot, which belongs to the category of Large Language Models (LLM), thas was developed by OpenAI^3^. These models are language models (i.e., a probabilistic distribution of word sequences) that rely on neural networks with millions of parameters. ChatGPT is trained on diverse and extensive data sources (e.g., books, websites, and so on) and exhibits a groundbreaking ability to generate relevant and context-aware responses, making it a promising tool in the medical education area. The training data cut-off was September 2021. This means that data after this date were not used for training. The model is defined by OpenAI as *Our fastest model, great for most everyday tasks*.

On the other hand, GPT-4, is a large multimodal (i.e., accepts text and image as input) model, also developed by OpenAI, faster, with more parameters and better performance than ChatGPT. More details and a comparison between both LLM used in this work are available in [27]. The model is defined by OpenAI as *Our most capable model, great for tasks that require creativity and advanced reasoning*. For the purpose of this study, we utilized the version of ChatGPT/GPT-4 that was released on May 3^rd^ 2023, as documented in OpenAI’s release notes [28]. This version was used to answer all the rheumatology questions from the MIR exams in our study.

### 2.2. MIR exam

The MIR is a competitive examination required for entry into specialist medical training in Spain. It is comprised of 210 questions from more than thirty competencies (i.e., pediatrics, hematology, ophthalmology), see the Supplementary Material Excel File “MIR Competencies”, and follows a multiple-choice format (i.e., since the 2015-2016 academic year, the number of choices decreased from five to four), with only one correct answer. Each question on the exam typically presents a clinical case or a factual query, and the exam also includes image-based questions. The number of questions per competency varies per year, and both the exam and the answers are officially published by the Spanish Ministry of Health [29]. The questions from this examination serve as an invaluable resource for this study, as they represent standardized, expertly crafted questions designed to evaluate a comprehensive understanding of medical subjects, and, therefore, are leveraged to evaluate the accuracy and clinical reasoning of ChatGPT and GPT-4 when exposing them to the rheumatology discipline.

### 2.3. Inclusion criteria

Rheumatology-related questions from MIR exams published from 2009-2010 to 2022-2023 were included in this study. Questions assigned to other specialities, (e.g., pediatrics, orthopedics), that intersected with the field of rheumatology were also included.

On the other hand, questions containing images were excluded from the analysis. This decision was taken because of the current limitations of ChatGPT, as it is primarily a text-based model and does not possess the capability to process or interpret image data. GPT-4 allows images as input data, but this feature was not publicly available at the time of this research. Hence, any questions in the MIR exams that were dependent on visual information, such as graphs, pictures, or clinical image data (e.g., x-ray), were not included. Finally, questions invalidated by the Spanish Ministry of Health were also excluded from the analysis.

### 2.4. Methodology

Questions from the MIR exams were used as prompts. For each question provided to ChatGPT/GPT-4, the sentence *“Justify your answer”* was added at the end. The responses generated by ChatGPT and GPT-4 to the rheumatology questions were evaluated by six independent rheumatologists. Three of them, ZRR, LCO, and CPR, in addition to being practising clinicians, are MIR training professors. The length of the text generated by the models was not artificially limited.

The medical experts evaluated the clinical reasoning of the chatbots followed in each of the responses. Their evaluation was based on a 1-5 scale, where a score of 5 indicates that the reasoning was entirely correct and flawless, while a score of 1 signifies that the reasoning was inconsistent or contained significant errors. Intermediate scores were used to denote minor errors in reasoning and the severity of these errors was reflected in the score; see Table 1. The final clinical reasoning score was assigned following a majority vote approach. In case of a tie, the worst score was chosen. The evaluators were also asked to justify the score given to ChatGPT/GPT-4 clinical reasoning when necessary. After that, each question was categorized based on the type of disease being asked about, and classified into factual or clinical case questions.

**Table 1.**
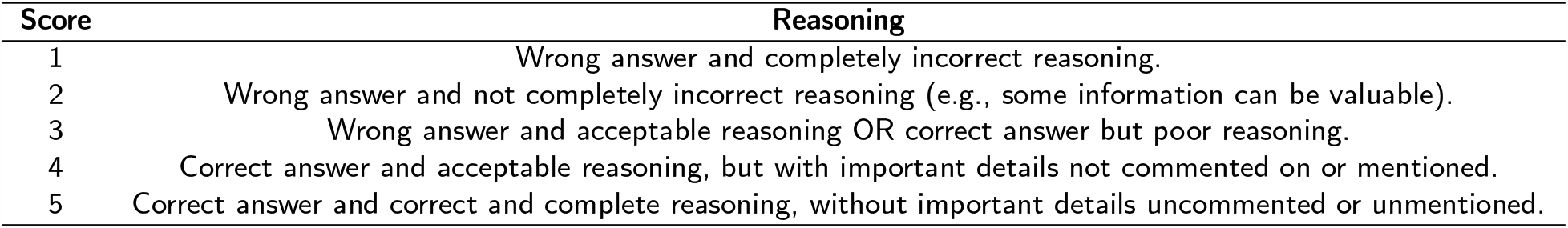
Template of the reasoning score given to the evaluators.

For this study, we solely relied on the initial responses generated by ChatGPT/GPT-4, without employing the *regenerate response* function. The questions were prompted in Spanish, exactly as they were extracted from the exam. Nonetheless, an English translation of both the question and the clinical reasoning by the LLM was obtained with DeepL and shown in the Supplementary Material Excel File “Data”, RheumaMIR [30]. In cases where the answers provided by the models were not singular, a new prompt was used to ask for a single and unique response with the following instruction: “*If you had to choose one of the answers, the most relevant one, which would it be?*”

The clinical reasoning followed by the models and evaluated by the medical experts was also evaluated in Spanish.

Finally, a questionnaire was provided to each evaluator to assess the overall performance of the language models and their suitability for use in education and clinical practice. This questionnaire comprises seven free-text questions and can be found in the Supplementary Material ”Questionnaire”.

#### 2.4.1. Variables

Two main variables were considered for evaluation:

- Accuracy, defined as the match between the official MIR question response and the chosen option by ChatGPT/GPT-4.
- Score assigned to the clinical reasoning of ChatGPT/GPT-4 by the six evaluators.

Covariables to be considered in this study included: year of the exam question, type of question, patient’s gender, pathology that the question addresses, and chatbot model used.

### 2.5. Statistical analysis

Dichotomous and categorical variables were summarized using proportions. Continuous variables were summarized using the median and the first and third quartiles (Q1–Q3). The distribution of correct answers (i.e., accuracy) among the different covariates was analyzed using chi-square or Fisher’s test, depending on the number of events. Differences between LLM, in terms of accuracy, were evaluated using McNemar’s test.

The degree of agreement in the score assigned to the clinical reasoning by the evaluators was analyzed using Krippendorff’s alpha coefficient and Kendall’s coefficient with and without tie correction coefficients [31]. The Kappa-Fleiss coefficient was not used as raters are considered unique (i.e., all evaluators justify the reasoning of all questions). The final clinical reasoning score given to each question was determined by a majority vote among evaluators. In the event of a tie, the worst score was chosen.

Differences between LLM, in terms of clinical reasoning, were evaluated using Wilcoxon signed-rank test. The effect of covariates on the clinical reasoning score was studied using ordinal logistic regression.

R version 4.3.1 was used to perform the statistical analysis.

### 2.6. Ethics board approval

As suggested in [10], the Hospital Clínico San Carlos (HCSC) Ethics Review Board evaluated this project, 23/370-E, and stated that this committee was not competent to evaluate studies of this type, since they do not encompass human subjects, or the use of biological samples, or personal data.

## 3. Results

The questions evaluated by ChatGPT/ GPT-4, the answer by both systems, as well as the official answer, the medical experts’ evaluation of the clinical reasoning, the year, genre, type of question, whether the question was invalidated or not, the type of disease being asked about, and the English translation of the questions and the clinical reasoning are shown in the Supplementary Material Excel File ’*Data*’. This dataset, called RheumaMIR, is also shared through Zenodo [30].

### 3.1. Description

After applying the inclusion criteria, 143 questions from 14 MIR exams remained (i.e., academic years 2009-2010 to 2022-2023). Table 2 shows the dataset characteristics. The median number of questions (Q1-Q3) per year is 11 (9.25-12) and the most prevalent disease being asked about is vasculitis. Most of the questions, 65.73%, were clinical cases, and the sex of the clinical case subjects was evenly distributed.

**Table 2.** Dataset description. ^±^: Including amyloidosis and autoinflammatory syndromes, fibromyalgia, igG4-related disease inflammatory myopathy, osteoarthritis, others, sarcoidosis, Sjögren’s syndrome.

| Variable | n (%) |
| --- | --- |
| **Year/number of questions** |  |
| 2009-2010 | 12 (8.39%) |
| 2010-2011 | 11 (7.69%) |
| 2011-2012 | 11 (7.69%) |
| 2012-2013 | 12 (8.39%) |
| 2013-2014 | 5 (3.50%) |
| 2014-2015 | 11 (7.69%) |
| 2015-2016 | 12 (8.39%) |
| 2016-2017 | 9 (6.29%) |
| 2017-2018 | 10 (6.99%) |
| 2018-2019 | 6 (4.20%) |
| 2019-2020 | 11 (7.69%) |
| 2020-2021 | 15 (10.49%) |
| 2021-2022 | 6 (4.20%) |
| 2022-2023 | 12 (8.39%) |
| **Disease** |  |
| Scleroderma | 8 (5.59%) |
| Microcrystalline arthritis | 9 (6.29%) |
| Infective arthritis | 11 (7.69%) |
| Rheumatoid arthritis | 12 (8.39%) |
| Spondyloarthropathies | 14 (9.79%) |
| Systemic lupus erythematosus | 16 (11.19%) |
| Bone metabolism | 20 (13.99%) |
| Vasculitis | 23 (16.08%) |
| Others<sup>±</sup> | 30 (20.98%) |
| **Type of question** |  |
| Factual | 49 (34.27%) |
| Clinical case | 94 (65.73%) |
| **Sex** |  |
| No sex/Newborn | 2 (1.40%) |
| Women | 42 (29.37%) |
| Not applicable | 49 (34.27%) |
| Men | 50 (34.97%) |

### 3.2. Accuracy

Out of 143 questions, GPT-4 correctly answered 134 (93.71%), demonstrating a high level of accuracy. On the other hand, ChatGPT accurately answered 95 questions (66.43%), indicating a somewhat lower level of performance in comparison to GPT-4 (i.e., McNemar’s test p-value=1.17×10^−09^).

ChatGPT did not correctly answer any of the questions that GPT-4 failed to answer correctly. Moreover, out of the nine questions that GPT-4 got wrong, seven of them also matched the answer given by ChatGPT. Table 3 shows the number and percentage of errors per model and covariate. Eventually, none of the covariates were associated with the number of errors, neither in GPT-4 nor in ChatGPT.

**Table 3.** Number of errors per model and covariate.

| Variable | ChatGPT<br>error | GPT-4<br>error |
| --- | --- | --- |
| **Year** |  |  |
| 2009-2010 | 3 (25.00%) | 1 (8.33%) |
| 2010-2011 | 4 (36.36%) | 2 (18.18%) |
| 2011-2012 | 4 (36.36%) | 1 (9.09%) |
| 2012-2013 | 3 (25.00%) | - |
| 2013-2014 | 2 (40.00%) | - |
| 2014-2015 | 3 (27.27%) | - |
| 2015-2016 | 4 (33.33%) | - |
| 2016-2017 | 3 (33.33%) | - |
| 2017-2018 | 3 (30.00%) | - |
| 2018-2019 | 4 (66.67%) | 2 (33.33%) |
| 2019-2020 | 4 (36.36%) | 1 (9.09%) |
| 2020-2021 | 6 (40.00%) | 2 (13.33%) |
| 2021-2022 | 1 (16.67%) | - |
| 2022-2023 | 4 (33.33%) | - |
| **p-value** | 0.979 | 0.233 |
| **Disease** |  |  |
| Scleroderma | 2 (25.00%) | 1 (12.50%) |
| Microcrystalline arthritis | 6 (66.67%) | - |
| Infective arthritis | 4 (36.36%) | - |
| Rheumatoid arthritis | 3 (25.00%) | 1 (8.33%) |
| Spondyloarthropathies | 2 (14.29%) | 1 (7.14%) |
| Systemic lupus erythematosus | 6 (37.50%) | - |
| Bone metabolism | 5 (25.00%) | 1 (5.00%) |
| Vasculitis | 10 (43.48%) | 3 (13.04%) |
| Others | 10 (33.33%) | 2 (6.67%) |
| **p-value** | 0.355 | 0.816 |
| **Type of question** |  |  |
| Factual | 18 (36.73%) | 6 (12.24%) |
| Clinical case | 30 (31.91%) | 3 (3.19%) |
| **p-value** | 0.580 | 0.063 |
| **Sex** |  |  |
| Women | 14 (33.33%) | 1 (2.38%) |
| Men | 15 (30.00%) | 2 (4.00%) |
| **p-value** | 1 | 0.823 |

### 3.3. Clinical reasoning

The Krippendorff’s alpha and Kendall’s coefficients with and without tie correction, for GPT-4 clinical reasoning, considering the six evaluators, were 0.225, 0.452 and 0.269, respectively. These values were higher when analyzing ChatGPT clinical reasoning, 0.624, 0.783 and 0.636, respectively. That is, the evaluators agreed more on the scores given to ChatGPT than on the scores given to GPT-4. A more detailed analysis of the main differences between evaluators’ scores can be found in the Supplementary Material File Section ’Evaluator Accordance’.

The median (Q1-Q3) value of the average score for the clinical reasoning of GPT-4 was 4.67 (4.5-4.83), while for ChatGPT was 4.5 (2.33-4.67). There exist statistically significant differences in the clinical reasoning score, after applying the majority vote, of both LLM (i.e., Wilcoxon signed rank test p-value=4.47×10^−9^).

Regarding the covariates, there were no statistically significant differences in the clinical reasoning score for GPT-4 / ChatGPT after applying ordinal logistic regression models. Figure 1 shows the proportion of scores given by the evaluators, grouped by score. With the exception of one evaluator, Evaluator 6, the most repeated score given was 5 for both models, with a small percentage of low scores (i.e., 1, 2, 3). The comparison of the scores given by the evaluators to the clinical reasoning of ChatGPT and GPT-4, grouped by evaluation, is shown in Supplementary Figure 1. For both figures, the majority vote statistics are shown.

**Figure 1:**
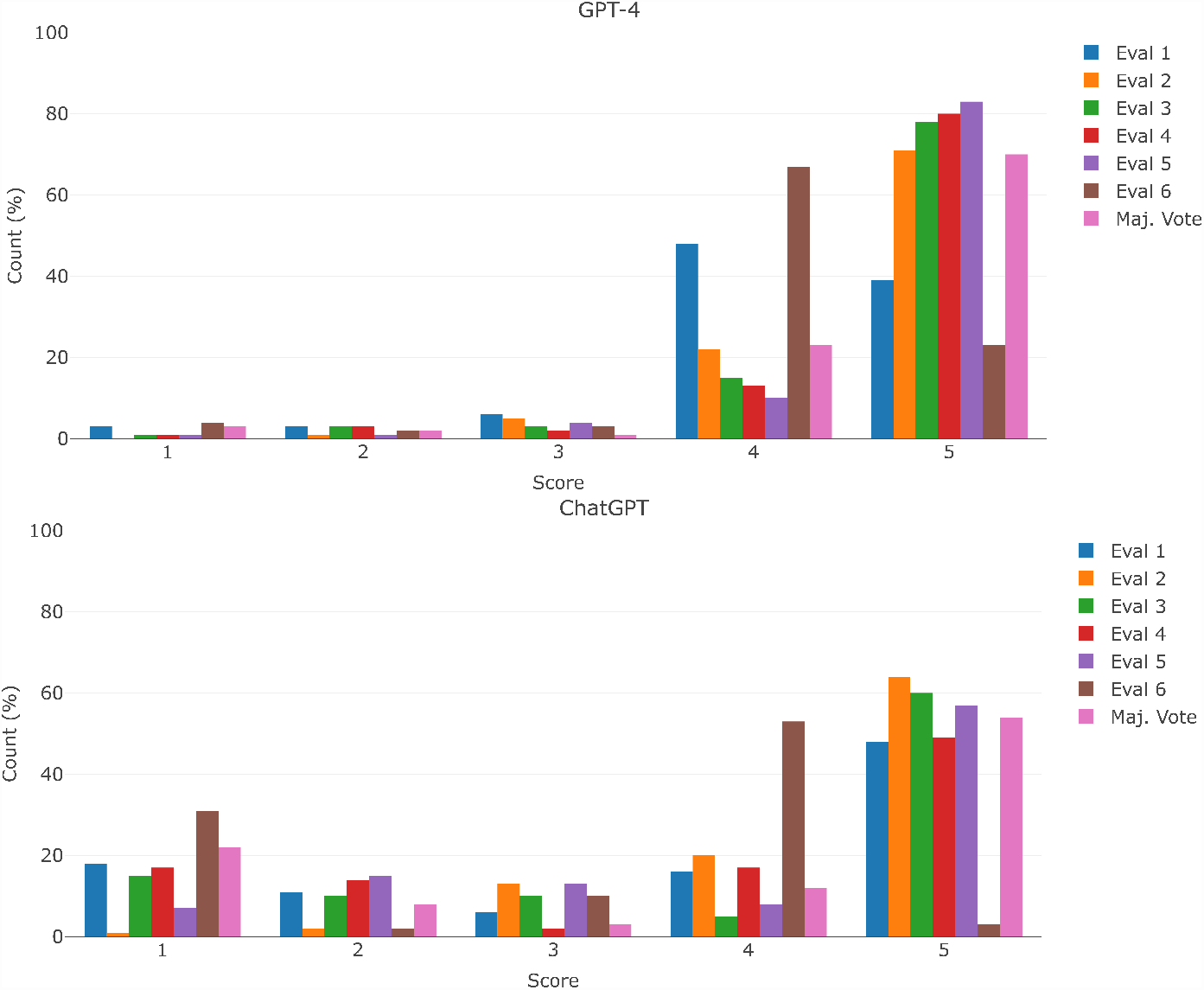
Distribution of the scores given by the evaluators

On its behalf, Figure 2 shows the proportion between the clinical reasoning score and the disease addressed in the questions. Supplementary Figures 2, 3 and 4, show the proportion between the clinical reasoning score and the year, the type of question, and the genre. There does not appear to be a clear trend between the reasoning score and the covariates shown in these plots. Finally, the completed questionnaires can be found in the Supplementary Material ’Questionnaire’. The evaluators concur on:

**Figure 2:**
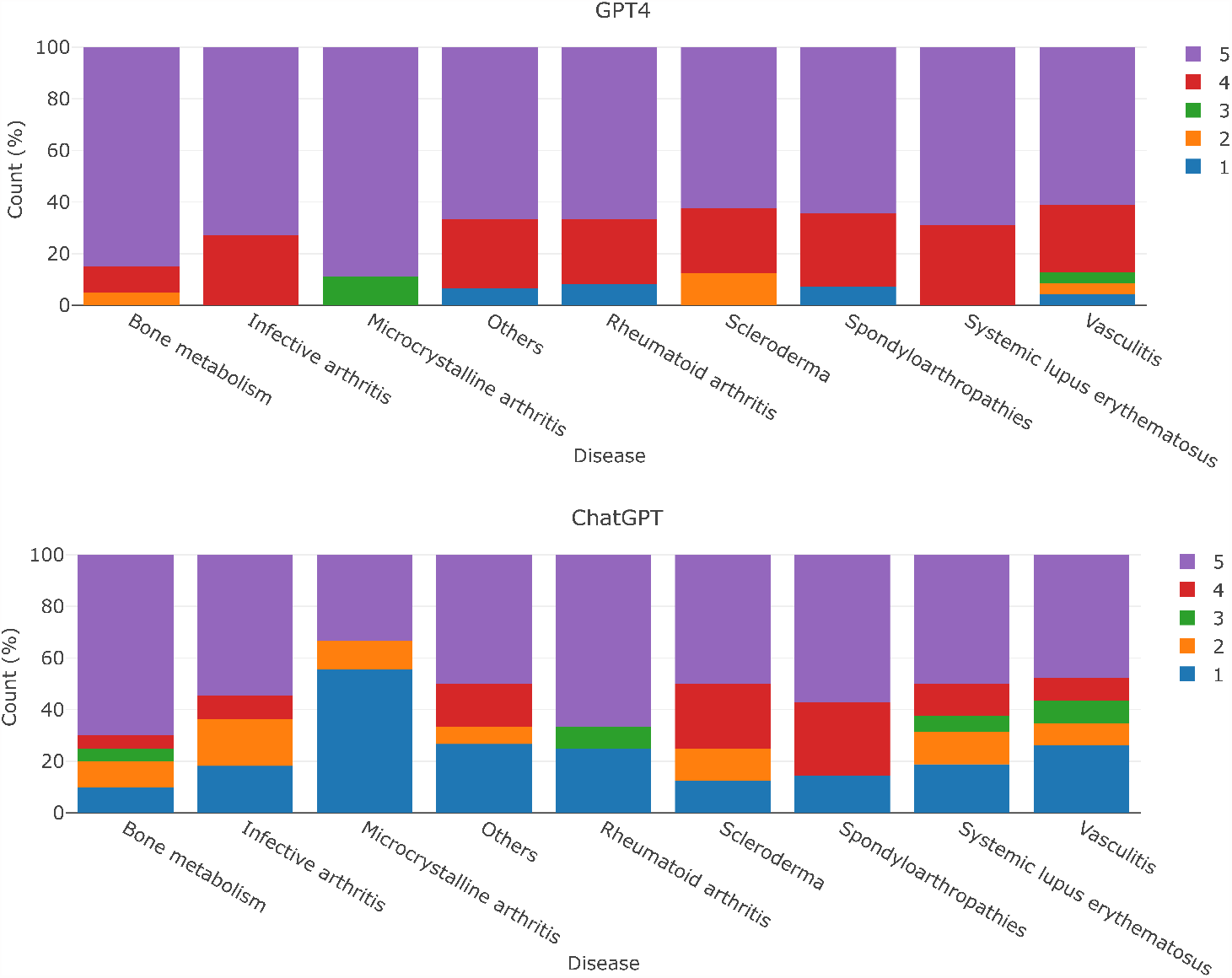
Clinical reasoning score according to the disease addressed in the question after taking the majority vote. In case of a tie, the worst score was chosen

- The potential usefulness of this tool, particularly in creating educational content, albeit under expert supervision.
- The language used in the responses may lack technical precision and could be suitable for students, but not in other scenarios (i.e., official medical documentation).
- ChatGPT/GPT-4 models are unaware of the limitations and scope of their knowledge, sometimes justifying facts in a tortuous manner and potentially misleading the reader.

## 4. Discussion

In this study, we have evaluated the accuracy and clinical reasoning of two LLM in answering rheumatology questions from Spanish official medical exams. To our knowledge, this is the first study to evaluate the usefulness of LLM applied to the training of medical students with a special focus on rheumatology.

The ability of GPT-4 to answer questions with high accuracy and sound clinical reasoning is remarkable. This could make such models valuable learning tools for medical students. However, ChatGPT/GPT-4 LLM are only the first models that have reached the general public in the rapidly expanding field of LLM chatbots. At present, a myriad of additional models are under development. Some of these nascent models are not only pre-trained in biomedical texts [32, 33], but are also specifically designed for a broad range of tasks (e.g., text summarization, question-answering and so on).

Studies with a similar objective to this one have been conducted. For example, a Spanish study [34], evaluated ChatGPT’s ability to answer questions from the 2022 MIR exam. In this cross-sectional and descriptive analysis, 210 questions from the exam were entered into the model. ChatGPT correctly answered 51.4% of the questions. This resulted in a 7,688 position, slightly below the median of the population tested but above the passing score.

In another research [35], the proficiency of ChatGPT in answering higher-order thinking questions related to medical biochemistry, including eleven competencies such as basic biochemistry, enzymes, chemistry and metabolism of carbohydrates, lipids and proteins, oncogenesis and immunity, was studied. Two-hundred questions were randomly chosen from an institution’s question bank and classified according to the Competency-Based Medical Education. The answers were evaluated by two expert biochemistry academicians on a scale of zero to five. ChatGPT obtained a median score of 4 out of 5, with oncogenesis and immunity competition having the lowest score and basic biochemistry the competition with the highest.

Research of a similar nature was conducted in [36]. In this study, the authors appraised the capability of ChatGPT in answering first- and second-order questions on microbiology (e.g., general microbiology and immunity, musculoskeletal system, skin and soft tissue infections, respiratory tract infections and so on) from the Competency Based Medical Education curriculum. A total of 96 essay questions were reviewed for content validity by an expert microbiologist. Subsequently, ChatGPT responses were evaluated on a scale of 1 to 5, with five being the highest score, by three microbiologists. A median score of 4.04 was achieved.

On the other hand, ChatGPT was tested on the Plastic Surgery In-Service examinations from 2018 to 2022 and its performance was compared to the national average performance of plastic surgery residents [37]. Out of 1,129 questions, ChatGPT answered 630 (55.8%) correctly. When compared with the performance of plastic surgery residents in 2022, ChatGPT ranked in the 49^th^ percentile for first-year residents,but its performance fell significantly among residents in higher years of training, dropping to the 0^th^ percentile for 5^th^ and 6^th^ -year residents.

Another study was conducted by researchers in [38], who aimed to assess whether ChatGPT could score equivalently to human candidates in a virtual objective structured clinical examinations in obstetrics and gynecology. Seven structured examination questions were selected, and the responses of ChatGPT were compared to the responses of two human candidates, and evaluated by fourteen qualified examiners. ChatGPT received an average score of 77.2%, while the average historical human score was 73.7%.

Moreover, the authors in [39] instructed ChatGPT to deliver concise answers to the 24-item diabetes knowledge questionnaire, consisting of a clear “*Yes*” or “*No*” response, followed by a concise rationale comprising two sentences for each question. The authors found that ChatGPT successfully answered all the questions.

In [40], the researchers were interested in evaluating the performance of ChatGPT on open-ended clinical reasoning questions. Therefore, fourteen multi-part cases were selected from clinical reasoning exams administered to first and second-year medical students, and provided to ChatGPT. Each case was comprised of 2-7 open-ended questions and was shown to ChatGPT twice. ChatGPT achieved or surpassed the pre-established passing score of 70% in 43% of the runs (12 out of 28), registering an average score of 69%.

Some studies showed remarkable performance, for instance, a research study evaluated the performance of ChatGPT in medical physiology university examination of phase I MBBS [41]. In this investigation, ChatGPT correctly answered 17 out of 20 multiple-choice questions, while providing a comprehensive explanation for each one. On their side, researchers in [42] proposed a four-grading system to classify the answers of ChatGPT, to note, comprehensive, correct but inadequate, mixed with correct and incorrect/outdated data, and completely incorrect. ChatGPT showed a 79% and a 74% of accuracy when answering questions related to cirrhosis and hepatocellular carcinoma. However, only the 47% and 41% of the answers were classified as comprehensive.

Conversely, in another research [43] in which ChatGPT was exposed to the family medicine course’s multiple-choice exam of Antwerp University, only 2/125 students performed worse than ChatGPT. Since the questions were prompted in Dutch language, the potential correlation between ChatGPT’s low performance and the proportion of Dutch texts used in its training could be a factor worth considering for this discordant result.

Another study, [44], evaluated ChatGPT’s performance on standardized admission tests in the United Kingdom, including the BioMedical Admissions Test (BMAT), Test of Mathematics for University Admission (TMUA), Law National Aptitude Test (LNAT), and Thinking Skills Assessment (TSA). A dataset of 509 multiple-choice questions from these exams, ranging from 2019 to 2022 was used. The results varied among specialities. For BMAT, the percentage of correct answers varied from 5% to 66%, for TMUA varied from 11% to 22%, for LNAT from 36% to 53%; and for TSA from 42% to 60%. The authors concluded that while ChatGPT demonstrated potential as a supplemental tool for areas assessing aptitude, problem-solving, critical thinking, and reading comprehension, it showed limitations in scientific and mathematical knowledge and applications.

The results shown by most of these studies are in line with our results, the average score of ChatGPT is between 4 and 5 (on a scale of five elements) when answering medical-related questions. However, in these studies, GPT-4 performance was not evaluated. On the basis of our results, we can postulate that there would be an increase in accuracy in comparison to those obtained by ChatGPT. In addition, to solve some of the limitations identified by our evaluators, such as the employment of a language that can lack technical precision by the models, LLM could perform better if trained or fine-tuned with biomedical texts.

A large part of the concerns and doubts that arise from using these models are due to regulatory and ethical issues. Some of the ethical dilemmas have been highlighted in [45]. For instance, the authors pointed out that LLM reflect any false, outdated, and biased data from which the model was trained, and that they could not reflect the latest guidelines. Other authors go further and declare that these types of models should not be used in clinical practice [46]. The motivation behind this statement lies in the presence of biases such as clinician bias, which may exacerbate racial-ethnic disparities, or hallucinations meaning that ChatGPT produces high levels of confidence in its output even when insufficient or masked information in the prompt. According to the authors, this phenomenon could lead users to place unwavering trust in the output of chatbots, even if it contains unreliable information. This was also pointed out by our evaluators, as shown in the Supplementary Material ’Questionnaire’. The firmness with which these models justify erroneous reasoning may limit their potential usefulness. Eventually, the authors also claimed that these models are susceptible to “Falsehood Mimicry”, that is, the model will attempt to generate an output that aligns with the user’s assumption rather than clarifying questions. In our study, due to the nature of the questions, we were unable to assess racial or ethnic disparities. However, we did not find any gender bias when considering the clinical case questions.

Regarding regulation, the arrival of these models has led to greater efforts being made to regulate the use of AI. The EU AI Act [47] is a good example of this. According to our results, in these early stages of LLM, the corpus used for training, as well as the content generated by them, should be carefully analyzed.

During the writing of this manuscript, new articles have emerged. For instance, in [48] authors studied the performance of ChatGPT when introducing the most searched keywords related to seven rheumatic and musculoskeletal diseases. The content of each answer was evaluated in terms of usefulness for patients with the ChatGPT in a scale from 1 to 7 by two raters.

Finally, further analysis is needed to explore these observations and understand their implications for the development and use of AI in medical education and practice.

### 4.1. Limitations

- Two chatbots were primarily used in this study, ChatGPT and GPT-4, both owned by OpenAI. However, other LLM such as BARD or Med-PaLM2 by Google, Claude 2 by Anthropic or LLaMA and LIMA by Meta are in development. Some of them are publicly available. To provide a better overview of other LLM, the accuracy of BARD (60.84%) and Claude 2 (79.72%) was calculated and compared against ChatGPT/GPT-4 in the Supplementary Material File Section ’LLM comparison’.
- The evaluators’ coefficient of agreement oscillates between 0.225 and 0.452 for the clinical reasoning of GPT-4, although the most repeated score of five out of six evaluators is 5 (the percentage of four and five scores oscillates between 87.41% and 93.70%), see Figure 1. This phenomenon is known as “Kappa paradoxes” (i.e., ’high agreement, but low reliability’), and tends to appear in skewed distributions such as the one presented in this study. More details can be found in [31]. In this study, since the ChatGPT clinical reasoning score distribution is less skewed, the reliability coefficient values are higher than the ones obtained with GPT-4.
- To ensure reproducibility and facilitate the comparison of results, each question could have been submitted twice to ChatGPT/GPT-4, a strategy supported by previous research endeavours, [42], and a default feature of BARD. However, in the tests we conducted, the responses were consistent across iterations and this would have doubled the workload of evaluators, so we chose to include more questions from a single run, rather than fewer questions run multiple times.
- The format of each question could have been transformed from multiple choice to open-ended. With this approach, it could have been possible to delve deeper into ChatGPT’s clinical reasoning. As explained in the previous point, this would have doubled the workload. Additionally, there are no open questions in the MIR exams.
- When conducting such studies, it is crucial to consider the evolution of knowledge over time. Evaluating older questions with models trained on recent data may reveal disparities compared to previously accepted and conventional beliefs. Therefore, accounting for this temporal aspect is essential to ensure the accuracy and relevance of the study findings. Moreover, although not extensively explored in this research, one of the key concepts when using LLM chatbots is what is known as the *prompt* or input text that is entered into the model. Depending on how well-defined the prompt is, the results can vary significantly. In this study, we tried to adhere closely to the official question while minimizing any unnecessary additional text.
- Another identified limitation of the study is the absence of medical student evaluation for the models’ clinical reasoning. This would have allowed us to determine whether students can recognize the main issues discussed above (e.g., bias, falsehood mimicry), and analyze to what extent these limitations may affect their usefulness.

### 4.2. Conclusion

The accuracy of ChatGPT and GPT-4 in answering the rheumatology questions of the Spanish access exam to specialized medical training is high, as well as the clinical reasoning. Nevertheless, discerning the veracity of such reasoning can pose a challenge for students in situations where the LLM experiences failures, given the comprehensive and seemingly accurate elaboration present in erroneous responses. However, these kinds of models hold the potential to serve as a valuable asset in the development of pedagogical materials, subject to rigorous expert evaluation.

## Supporting information

Supplementary Material 'Questionnaire'

Supplementary Excel File "MIR Competencies"

## CRediT authorship contribution statement

**Alfredo Madrid-García:** Conceptualization of this study, methodology, review, writing (original draft preparation). **Zulema Rosales-Rosado:** Evaluation. **Dalifer Freites-Nuñez:** Evaluation. **Inés Pérez-Sancristobal:** Evaluation. **Esperanza Pato-Cour:** Evaluation. **Chamaida Plasencia-Rodríguez:** Evaluation. **Luis Cabeza-Osorio:** Evaluation. **Leticia León-Mateos:** Methodology. **Lydia Abasolo-Alcázar:** Methodology. **Benjamín Fernández-Gutiérrez:** Conceptualization of this study. **Luis Rodríguez-Rodríguez:** Conceptualization of this study, methodology, review.

All of the authors were involved in the drafting and/or revising of the manuscript.

ChatGPT May 12^th^ version, 2023, (OpenAI, San Francisco, CA, USA) was utilized as a writing aid in the composition of this scientific article.

## Data availability statement

The data that support the findings of this study are openly available in Zenodo at http://doi.org/10.5281/zenodo.8153291.

## Footnotes

3 https://openai.com/

