## Supplementary Material 'Questionnaire' for "Harnessing ChatGPT and GPT-4 for Evaluating the Rheumatology Questions of the Spanish Access Exam to Specialized Medical Training"

### Supplementary Material: "Questionnaire for evaluating the use of language models in rheumatology"

**1.	Based on the clinical reasoning you have evaluated; do you think that the use of ChatGPT/GPT-4 could be useful as a content generator for the medical student who is preparing rheumatology-related content for the MIR exam?**

**Evaluator 1:** Yes, it could, but with supervision of the content by a subject specialist as most of the answers need "polishing".

**Evaluator 2:** I think ChatGPT-4 is useful as a content generator for the medical student. I think it is useful after the basic training rounds, I consider it valuable in the final stage of the student's training. Its supervised use with the subject trainers is recommended. Regarding ChatGPT as a content generator, it requires more guidance/supervision in its use.

**Evaluator 3:** I think so, as long as, it was supervised by a specialist in the subject or a trainer/teacher of this type of exams, but of course, as a method for practicing, I think it would be useful.

**Evaluator 4:** I think it could be useful as a tool to help generate content, but always with supervision of the content by a specialist in the field. For me it would be useful to use GPT-4, I would not use ChatGPT, in some cases to consult something and with a lot of supervision.

**Evaluator 5:** Yes, although it is true that it lacks training and supervision, especially GPT-4 can be useful.

**Evaluator 6:** Based on the hit rate it would be possible to use GPT-4 in this sense, not so much ChatGPT.

**2. Do you consider that the language used by both systems is appropriate for a medical student, does it contain/lack technicalities?**

**Evaluator 1:** It is not bad, for a MIR candidate it may be adequate, although I find it excessively colloquial, and it lacks technicalities. However, the answers are very well understood.

**Evaluator 2:** The language used in the ChatGPT/GPT-4 seems to me to be correct and appropriate for a medical student. The technicality of the content is just right to get the message across.

**Evaluator 3:** Sometimes it can be too colloquial, but in general, I think it is adequate for a MIR candidate, the explanations are usually very well understood.

**Evaluator 4:** It seems to me to be appropriate language for a recently finished doctor who is going to take an MIR exam. It is possible that the ChatGPT uses somewhat colloquial language.

**Evaluator 5:** Although some answers may be lacking in technicalities, the answers were well understood and for a MIR candidate the level could be adequate.

**Evaluator 6:** No, the language is not very academic for medical students.

**3. Are the justifications/reasoning of the ChatGPT/GPT-4 models clear, straightforward, and understandable?**

**Evaluator 1:** Chat-GPT's justifications are perhaps longer and more complete, but it fails more often so it is not applicable; GPT-4 gives more scientifically sound answers and is perhaps less broad, more direct.

**Evaluator 2:** In general, the justifications and reasoning in both models are clear and understandable. I consider ChatGPT-4 to be more direct in content.

**Evaluator 3:** Apart from a few cases, which could have been a little too concise and particularly convoluted, in general, I found the explanations to be well developed and well structured.

In particular, I felt that GPT-4 developed them better from a scientific point of view.

**Evaluator 4:** I find them understandable in 95% of cases or more, they are well structured and the documentation they use seems to be correct. Sometimes the justifications are a bit lengthy.

**Evaluator 5:** In general, although both systems give long justifications, my impression is that the GPT-4 ones are more scientifically sound and somewhat more straightforward and understandable than the ChatGPT ones.

**Evaluator 6:** The justifications are quite well-reasoned, even at great length in many cases.

**4. Do ChatGPT/GPT-4 models show awareness of the limitations and scope of their knowledge, avoiding speculation or incorrect answers when there is insufficient information?**

**Evaluator 1:** Not much, they always try to justify their answer (often incorrect) with whatever, without any criteria. In their favor, it must be said that at the end they always put the phrase that it is necessary to consult a doctor and that it is necessary to assess each individual case.

**Evaluator 2:** Not entirely, sometimes the content of the justifications/answers lacks reflection or debate. We found some questions with correct answers, not entirely well developed in their justification. The opposite is also observed (ChatGPT), where an incorrect answer is generated and the reasoning contains erroneous information as if it were correct. Students should approach their answers with caution.

**Evaluator 3:** Not always, sometimes, especially in the case of ChatGPT, incorrect answers are given and yet attempts are made to justify and explain even when the criteria on which they are based are not properly documented or up to date.

**Evaluator 4:** No, especially ChatGPT. It gives wrong answers and justifies them with wrong information as if they were correct.

**Evaluator 5:** Above all, GPT-4 is the system that manages to get it most right and to make the most accurate reasoning in my opinion.

**Evaluator 6:** Mostly they give an answer based on their knowledge, which may be wrong in some cases.

**5.	As a rheumatology specialist, what application do you think ChatGPT/GPT4 could have, or what use would you make of it as a practitioner?**

**Evaluator 1:** They seem to me to be tools that can make it easier and quicker to create teaching material, but they need the supervision of a professional to check both the content and the language.

**Evaluator 2:** I consider it useful as an additional tool for the medical specialist. It could be applied in medical diagnosis, for reading medical records, analysing symptoms, analysis and imaging tests and for educational purposes. With the eventual development of the tool, GPT-4 has the potential to transform medical care and improve patient care. However, its implementation should be treated with caution, taking into account ethical, legal and practical issues.

**Evaluator 3:** Mainly, I think they could be used on the one hand, for teaching (creation of materials, guides, help with exams) and, on the other hand, it can be a tool that can help in specific clinical doubts as long as we are aware that it would only be a first approximation, that is, accessing the scientific information available in a faster and more concise way (similar to an "Uptodate") but always being the content supervised and taking into account by the specialists, its scope and limitations.

**Evaluator 4:** I think it could be useful as a tool to help in clinical doubts for a first approach, to supervise the response and to access scientific information in a more targeted way. It can also help to create scientific and teaching material but always with supervision.

**Evaluator 5:** As a professional, I believe that these systems can help us to speed up the creation of teaching material (for classes, exams, gamification...) but with the supervision of a professional who reviews the content, which is crucial at this time.

**Evaluator 6:** GPT4 is much more accurate and could be useful for generating questions and evaluations for courses/lectures/sessions.

**6. In general terms, what weaknesses/limitations have you identified in the answers provided by ChatGPT/GPT-4 (maximum of 3)?**

**Evaluator 1:**

- Their reasoning is different from that of teachers and clinicians, which causes them to fail the most difficult questions because they often need exam technique to answer them or more in-depth reasoning.
- It seems that the sources from which they take the information are not very scientific, no reference is made in any case to EULAR/ACR...
- They justify the unjustifiable with a reasoning that you could come to believe, even if it is wrong, that is worrying.

**Evaluator 2:**

- It seems to me a limitation not to acknowledge the source or bibliographic reference of the justification or reasoning. The model may generate answers based on generic knowledge rather than specific medical protocols or classification criteria, for example.
- From a training point of view, medical students preparing for the MIR exam, in its use there should be a process in which experts can review the content.

**Evaluator 3:**

- Sometimes explanations too convoluted and not very concise and brief.
- Explanations sometimes too informal, particularly ChatGPT
- Outdated information and not based on adequate sources (ACR, EULAR, rheumatology books, etc...).

**Evaluator 4:**

- Lacks awareness of errors and justifies them as correct (especially ChatGPT).
- Language is sometimes unscientific, more so in ChatGPT, and does not seem to base answers on established classification criteria.

**Evaluator 5:** In general, I have found that exceptions or difficult cases are more likely to escape ChatGPT than GPT-4. That is, cases that fall outside the norm are more difficult to discern the appropriate response.

**Evaluator 6:**

- Unscientific language in some cases
- Excessively long and exhaustive explanations

**7. In general terms, what strengths have you identified in the answers provided by ChatGPT/GPT-4 (maximum of 3)?**

**Evaluator 1:** They provide a lot of information quickly and with training could probably be very useful at this time for the general public rather than the specialist.

**Evaluator 2:**

- The answers, generated quickly, produce a well-structured text.
- The language used is correct and understandable.

**Evaluator 3:**

- Speed and convenience
- Ease of access to information, especially better with GPT-4 which has more hits and better reasoning of options.
- Understandable language.

**Evaluator 4:** More strengths with the GPT-4 which seems "more scientific" and gets most of the questions right, few errors. The language it uses is understandable.

**Evaluator 5:**

- Chat-GPT: reasons between true and false in general, understandable language
- Chat-GPT4: more hits, understandable, reasoning between true and false in general

**Evaluator 6:**

- Very high percentage of correct answers to the questions.
- Up-to-date bibliography
