## Supplementary Material 'File' for "Harnessing ChatGPT and GPT-4 for Evaluating the Rheumatology Questions of the Spanish Access Exam to Specialized Medical Training"

Supplementary Figures

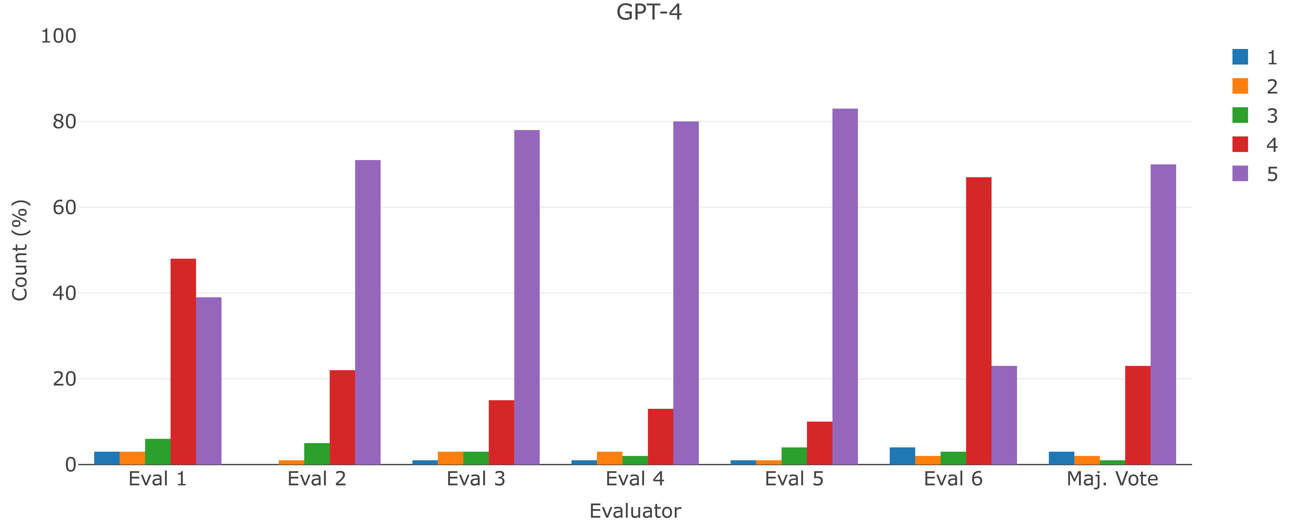

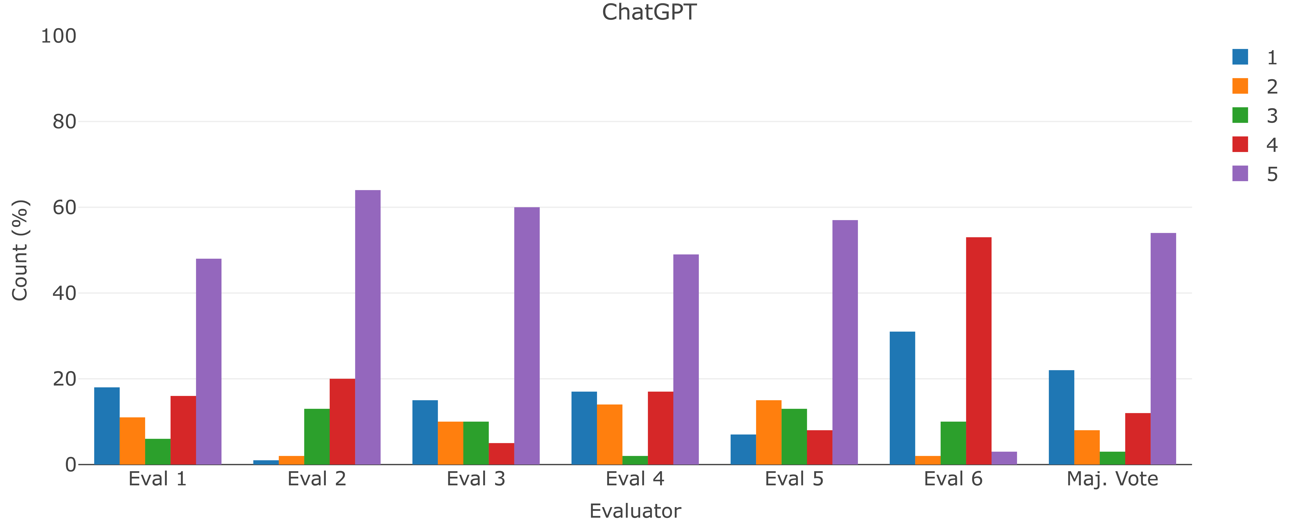

Supplementary Figure 1: Scores given by the evaluators to the clinical reasoning

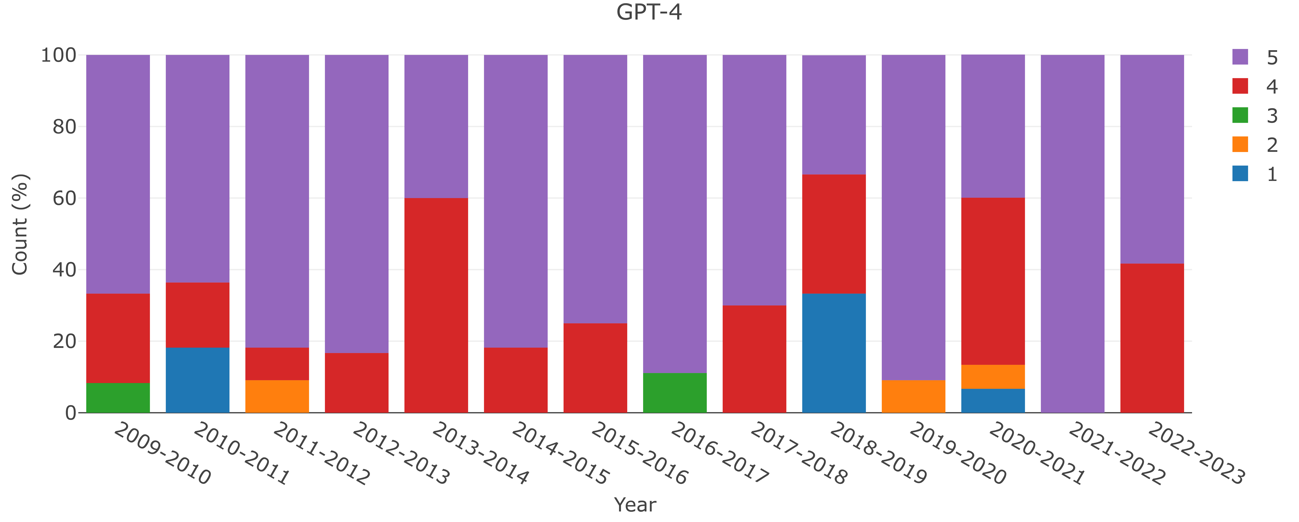

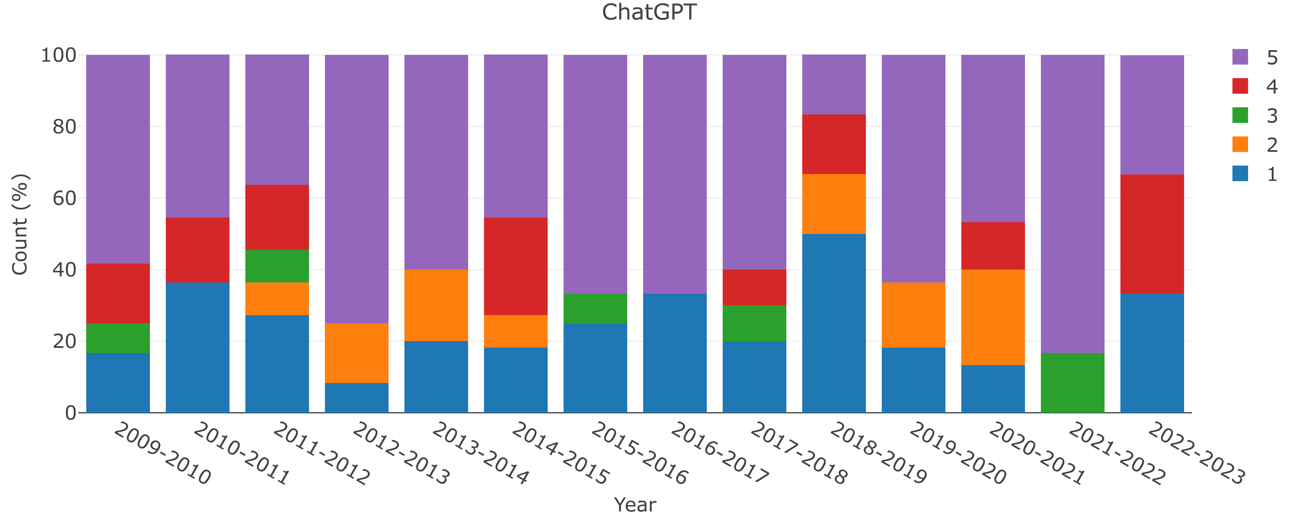

Supplementary Figure 2: Clinical reasoning score according to the year of the exam after taking the majority vote. In case of a tie, the worst score was chosen.

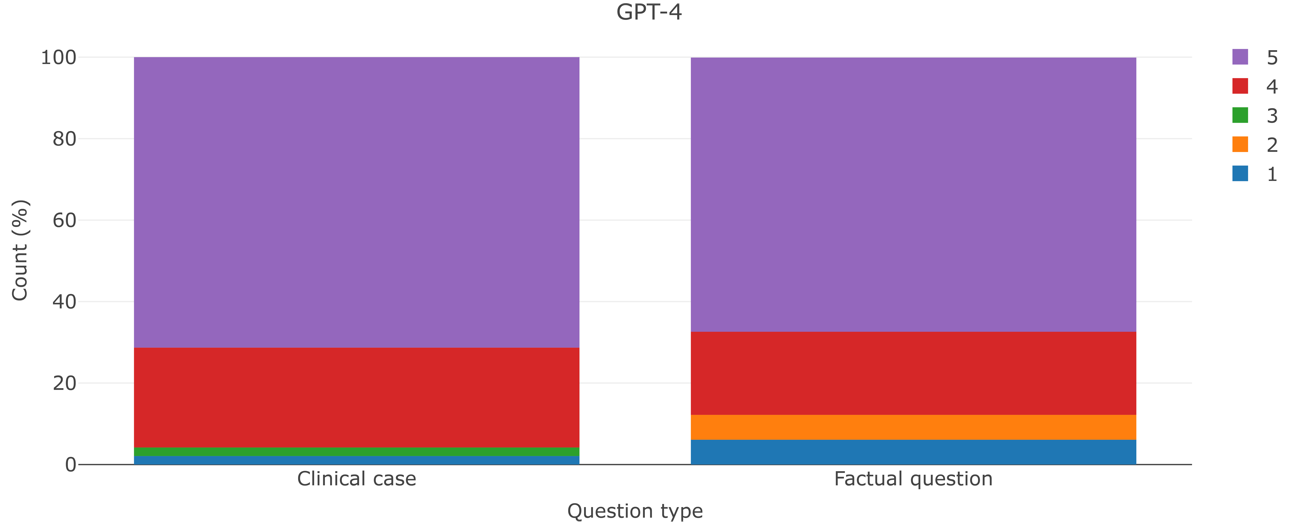

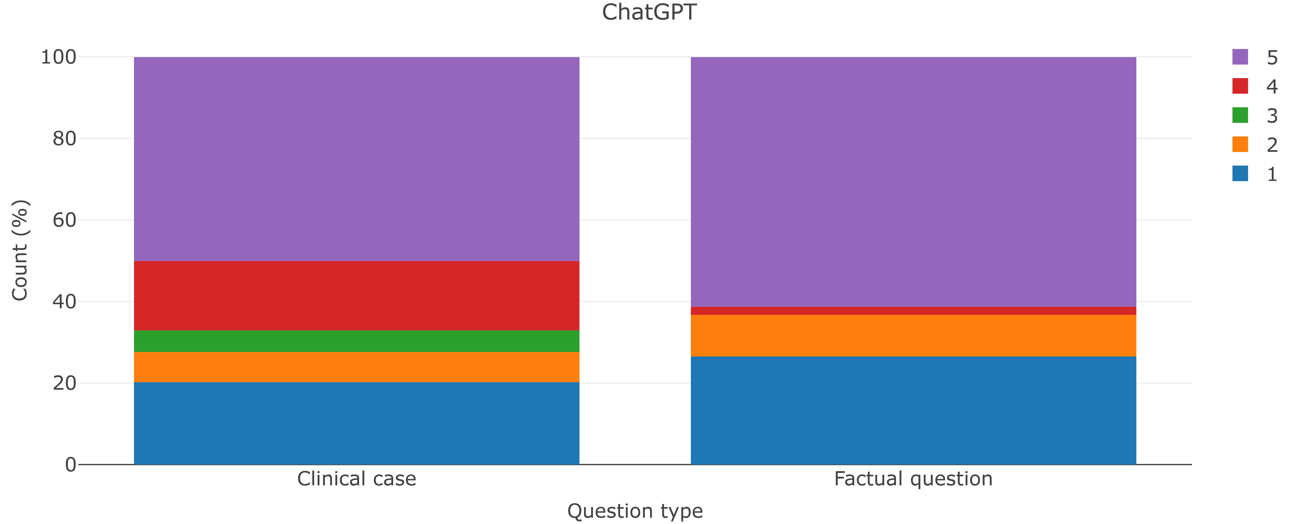

Supplementary Figure 3: Clinical reasoning score according to the type of the question after taking the majority vote. In case of a tie, the worst score was chosen

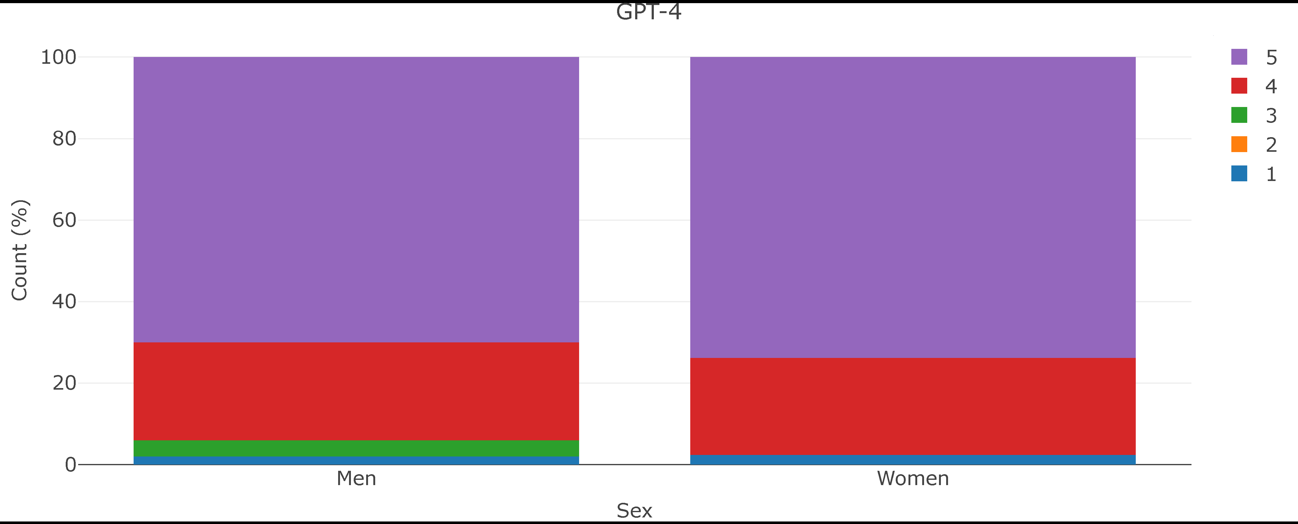

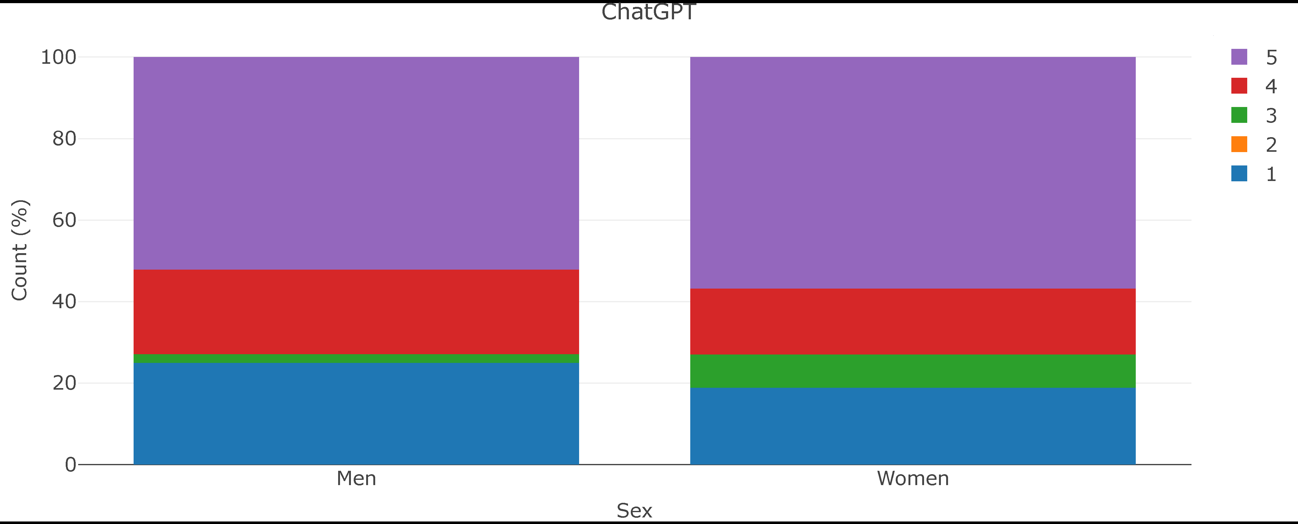

Supplementary Figure 4: Clinical reasoning score according to the sex after taking the majority vote. In case of a tie, the worst score was chosen

Evaluator accordance

Supplementary Table 1 shows the number of questions with two or more points (positive) of difference in the clinical reasoning of GPT-4 among evaluators. Supplementary Table 2 shows the number of questions with two or more point (negative) of difference. Supplementary Table 3 and 4 behave in the same way as Supplementary Tables 1 and 2 but with the ChatGPT clinical reasoning.

Supplementary Table 1: -2 points difference in the clinical reasoning GPT-4

| **Evaluator** | **1** | **2** | **3** | **4** | **5** | **6** |
| --- | --- | --- | --- | --- | --- | --- |
| **1** |  | 11 | 3 | 6 | 8 | 3 |
| **2** |  |  | 2 | 3 | 2 | 1 |
| **3** |  |  |  | 1 | 3 | 1 |
| **4** |  |  |  |  | 1 | 0 |
| **5** |  |  |  |  |  | 0 |
| **6** |  |  |  |  |  |  |

Supplementary Table 2: +2 points difference in the clinical reasoning GPT-4

| **Evaluator** | **1** | **2** | **3** | **4** | **5** | **6** |
| --- | --- | --- | --- | --- | --- | --- |
| **1** |  | 1 | 0 | 0 | 1 | 2 |
| **2** |  |  | 1 | 3 | 3 | 7 |
| **3** |  |  |  | 1 | 3 | 1 |
| **4** |  |  |  |  | 1 | 5 |
| **5** |  |  |  |  |  | 7 |
| **6** |  |  |  |  |  |  |

Supplementary Table 3: -2 points difference in the clinical reasoning ChatGPT

| **Evaluator** | **1** | **2** | **3** | **4** | **5** | **6** |
| --- | --- | --- | --- | --- | --- | --- |
| **1** |  | 36 | 5 | 5 | 11 | 1 |
| **2** |  |  | 2 | 0 | 1 | 2 |
| **3** |  |  |  | 4 | 10 | 1 |
| **4** |  |  |  |  | 9 | 2 |
| **5** |  |  |  |  |  | 2 |
| **6** |  |  |  |  |  |  |

Supplementary Table 4: +2 points difference in the clinical reasoning ChatGPT

| **Evaluator** | **1** | **2** | **3** | **4** | **5** | **6** |
| --- | --- | --- | --- | --- | --- | --- |
| **1** |  | 0 | 1 | 4 | 2 | 17 |
| **2** |  |  | 31 | 31 | 25 | 55 |
| **3** |  |  |  | 6 | 4 | 24 |
| **4** |  |  |  |  | 3 | 17 |
| **5** |  |  |  |  |  | 28 |
| **6** |  |  |  |  |  |  |

LLM comparison

The accuracy of BARD by Google, version 11^th^ July 2023, and Claude 2 by Antrophic, version 11^th^ July 2023, was compared against GPT-4 and ChatGPT. Only the accuracy of these two models were measured and not their clinical reasoning.

As it was done with the OpenAI solutions, the prompts were introduced in Spanish, exactly as we did for ChatGPT and GPT-4. Supplementary Figure 1 shows a comparison of the accuracy of all the LLM tested in this work.

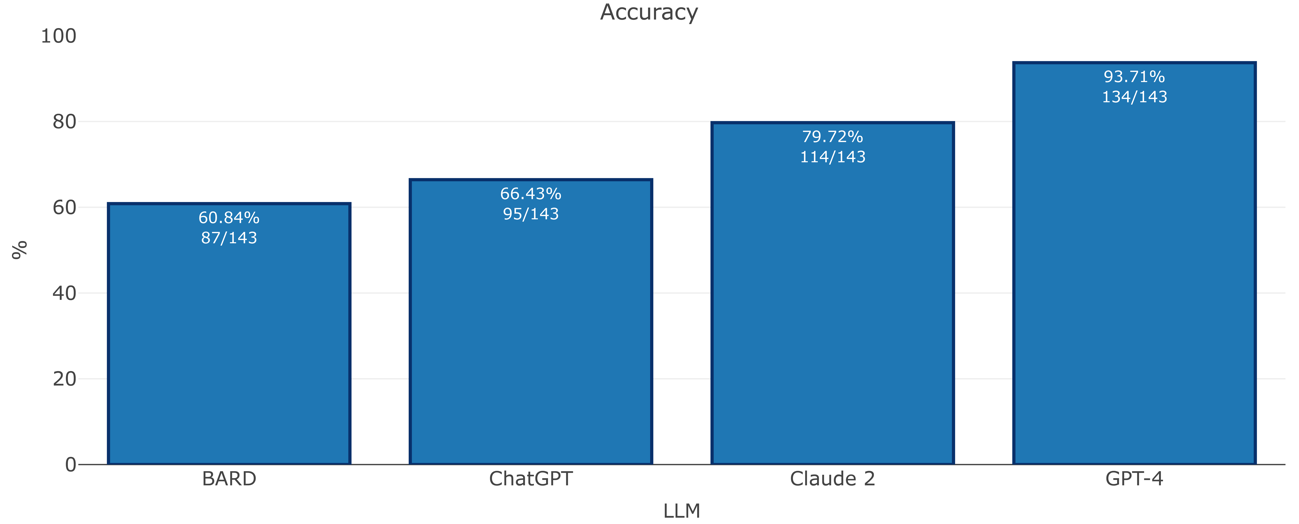

Supplementary Figure 5: Accuracy of BARD, ChatGPT, Claude 2 and GPT-4 in a dataset of 143 rheumatology questions
